# Immunogenicity of BNT162b2 Vaccine in Patients with Inflammatory Bowel Disease on Infliximab Combination Therapy: A Multicenter Prospective Study

**DOI:** 10.1101/2021.10.20.21265239

**Authors:** Mohammad Shehab, Mohamed Abu-Farha, Fatema Alrashed, Ahmad Alfadhli, Khazna AlOtaibi, Abdulla AlSahli, Thangavel Alphonse Thanaraj, Arshad Channanath, Hamad Ali, Jehad Abubaker, Fahd Al-Mulla

**Author notes:** **Corresponding authors:** Mohammad Shehab, Division of Gastroenterology, Department of Internal Medicine, Mubarak Alkabeer University Hospital, Kuwait University, Aljabreyah, Kuwait. And Fahd Almulla, Department of Genetics and Bioinformatics, Dasman Diabetes Institute (DDI), Dasman, Kuwait.

## Abstract

**Background:** Vaccination is a promising strategy to protect vulnerable groups like inflammatory bowel disease (IBD) patients against COVID-19 and associated severe outcomes. COVID-19 Vaccines clinical trials excluded IBD patients taking infliximab with azathioprine or 6-mercaptopurine (infliximab combination). Therefore, we sought to evaluate serologic responses to COVID-19 vaccination with the mRNA vaccine, BNT162b2 in IBD patients receiving infliximab combination therapy compared to healthy participants.

**Methods:** This is a multicenter prospective study. IBD patients were recruited at the time of attendance at infusion center between August 1^st^, 2021, and September 15^th^, 2021. Our primary outcome was the concentrations of SARS-CoV-2 antibodies 4-10 weeks after vaccination with two doses of BNT162b2 vaccine in IBD patients taking infliximab combination therapy (study group) compared to healthy participants group (control group). Both study and healthy participants groups were matched for age, sex and time-since-last-vaccine-dose using optimal pair matching method.

**Results:** In total 116 participants were recruited in the study, 58 patients in the study group and 58 in the control group. Median (IQR) IgG concentrations were lower in the study group [99 BAU/mL (40, 177)] than the control group [139 BAU/mL (120, 188)], following vaccination (p = 0.0032). Neutralizing antibodies was also lower in the study group compared to the control group [64% (23, 94) vs 91% (85, 94), p <0.001]. The median IgA levels in the study group was also significantly lower when compared to the control group [6 U/ml (3, 34) vs 13 U/ml (7, 30), p =0.0097]. In the study group, the percentage of patients who achieved positive IgG, neutralizing antibody and IgA levels were 81%, 75% and 40% respectively. In the control group, all participants (100%) had positive IgG and neutralizing antibody levels while 62% had positive IgA levels.

**Conclusion:** In patients with IBD receiving infliximab combination therapy, IgG, IgA and neutralizing antibody levels after BNT162b2 vaccine were lower compared to healthy participants. However, most patients treated with infliximab combination therapy seroconverted after two doses of the vaccine.

## Introduction

Inflammatory bowel disease (IBD) is an immune-mediated chronic inflammatory disease. The health of patients with IBD during the coronavirus disease 2019 (COVID-19) pandemic have been an area of concern due to increased susceptibility to infections.^1,2^ As with other immune-mediated inflammatory diseases, patients with IBD may require immunosuppressive drugs such as corticosteroids, thiopurines, tumor necrosis factor inhibitors (anti-TNFs), integrin receptor antagonists, anti-IL/12/23 inhibitors, and Janus kinase (JAK) inhibitor to achieve and maintain disease response and remission. The use of such medications has raised concerns regarding the increased susceptibility to acute respiratory syndrome coronavirus 2 (SARS-CoV-2) infection. Indeed, recent studies have confirmed association of corticosteroids and severe COVID-19 outcomes such as hospitalization and death.^3–6^ Therefore, expert consensus advocates that IBD patients should be vaccinated against SARS-CoV-2.^7,8^

The goal of vaccination is to produce a long-term immunity against infection, and this can be achieved, in part, by production of pathogen-specific antibody. Immunosuppressive drugs may reduce the effectiveness of some vaccines. Studies showed that response to pneumococcal^9^, influenza^10^, hepatitis A^11^ and B^12^ vaccination in patients with IBD receiving immunosuppressive agents is diminished compared with that in control individuals. However, the impact of IBD medications on COVID-19 vaccine efficacy is unknown. CLARITY study showed that infliximab is associated with attenuated serological responses to SARS-CoV-2 that were further blunted by immunomodulators used as combination therapy.^13^ However, to our knowledge, there is no study examined the impact of infliximab use with azathioprine or 6-mercaptopurine (infliximab combination) on COVID-19 vaccine efficacy compared to healthy individuals. The objective of this study is to evaluate serologic responses to COVID-19 vaccination with the mRNA vaccine, BNT162b2 (Pfizer/BioNTech) in patients with IBD receiving infliximab combination therapy compared to healthy participants.

## Method

### Recruitment and eligibility

A prospective multi center cohort study was conducted at two tertiary care centers. Patients were recruited at the time of attendance at the infusion centers between August 1^st^, 2021, and September 15^th^, 2021. Patients were eligible to be included if they : 1) had confirmed diagnosis of inflammatory bowel disease before the start of the study 2) were receiving infliximab with azathioprine or 6-mercaptopurine for at least 6 weeks or more for induction of remission or with at least one dose of drug received for maintenance of remission in the previous 8 weeks 3) have received two-dose of COVID-19 vaccination with BNT162b2 vaccine, 3 weeks apart, within 4-10 weeks before recruitment 4) at least are 18 years of age or older. Patients were excluded if they only received a single dose of the vaccine or if they were infected or had symptoms of SARS-CoV-2 previously since the start of the pandemic. In addition, patients who received other vaccines than the BNT162b2 were excluded. Patients who were received corticosteroids two weeks before the first dose of the vaccine up to the time of recruitment were also excluded. Finally, patients taking other immunomodulators such as methotrexate were also excluded. This study was performed and reported in accordance with Strengthening the Reporting of Observational Studies in Epidemiology (STROBE) guideline.^14^

Diagnosis of inflammatory bowel disease (IBD) was made according to the international classification of diseases (ICD-10 version:2016). Patients were considered to have IBD when they had ICD-10 K50, K50.1, K50.8, K50.9 corresponding to Crohn’s disease (CD) and ICD-10 K51, k51.0, k51.2, k51.3, k51.5, k51.8, k51.9 corresponding to ulcerative colitis (UC).^15^ In the study group, data regarding type and extent of IBD as well as duration of infliximab combination therapy were obtained.

Healthy Participants (control) group were individuals with no history of chronic medical illnesses such as diabetes, hypertension, cardiovascular disease, autoimmune diseases, osteoarthritis, chronic obstructive pulmonary disease, renal disease, asthma, hyperlipidemia, history of stroke and bleeding disorder.

### Outcome measures

Our primary outcome was the concentrations of SARS-CoV-2 antibodies including Immunoglobulin G (IgG), Immunoglobulin A (IgA) and neutralizing antibodies 4–10 weeks after vaccination with two doses of the BNT162b2 in patients with IBD receiving infliximab combination therapy (study group) compared with healthy participants (control group).

### Laboratory Methods

In this study, plasma levels of SARS-CoV-2-specific IgG and IgA antibodies were measured by enzyme-linked immunosorbent assay (ELISA) kit (SERION ELISA agile SARS-CoV-2 IgG and IgA SERION Diagnostics, Wu□rzburg, Germany) based on the manufacturers protocol. Units of IgG levels were reported as binding antibody units (BAU)/mL. values of below 31.5 BAU/mL were considered negative or nonprotective. The IgA levels were reported as Arbitrary Units (AU)/mL, values below 10 AU/mL were considered negative or nonprotective. Neutralizing antibody levels below 20% were considered negative or nonprotective. The positive and negative thresholds were determined as per manufacturer’s instructions. Results were construed by calculating inhibition rates for samples as per the following equation: Inhibition = (1 - O.D. value of sample/O.D. value of negative control) ×100%.

### Ethical consideration

This study was reviewed and approved by the Ethical Review Board of Dasman institute “Protocol # RA HM-2021-008” as per the updated guidelines of the Declaration of Helsinki (64th WMA General Assembly, Fortaleza, Brazil, October 2013) and of the US Federal Policy for the Protection of Human Subjects. The study was also approved by the Ministry of Health of Kuwait (reference: 3799, protocol number 1729/2021). Subsequently, patient informed written consent was obtained before inclusion in the study.

### Statistics

We performed descriptive statistics to characterize the study and control group. Standard descriptive statistics were used to present the demographic characteristics of patients included in this study and their measured antibody levels. Analysis was conducted in R (R Core Team, 2017). Data are expressed as medians with interquartile range (IQR) unless otherwise indicated. Categorical variables were compared using the Fisher’s exact test or Pearson’s Chi-squared test, and continuous variables were compared with the Kruskal–Wallis rank sum test or Wilcoxon rank sum test. P-value of less than or equal to 0.05 considered statistically significant. Both Infliximab combination therapy and healthy participants groups were matched for age, sex and time-since-last-vaccine-dose using Optimal pair matching method. The technique attempts to choose matches that collectively optimize an overall criterion. The criterion used was the sum of the absolute pair distances in the matched sample. In addition, percentage of positive IgG, neutralizing antibody and IgA levels was calculated in both groups.

## Results

### Baseline Cohort Characteristics

Patients were recruited between August 1^st^, 2021, until September 15^th^, 2021. In total, serology assays to quantify SARS-CoV-2 antibody levels were performed for 116 patients. The number of patients included in both the study group and control group was 58. Mean age was 33 in both groups and body mass index was lower in the study group compared to healthy participants (24.8 vs 26 kg/m^2^). Most patients in the study group had Crohn’s disease (60%) and 20% were smokers. The mean duration between the serology test and last dose of vaccine was 7 (±2) weeks. And median duration of infliximab combination therapy was 12 months (See table 1)

**Table 1:** Baseline characteristics of participants.

| **Variable** | **Study Group\* (n=58)** | **Control group (n=58)** |
| --- | --- | --- |
| Mean age (years) | 33.2 | 33 |
| Sex n (%) |  |  |
| Male | 33 (56%) | 31(53%) |
| Female | 25 (44%) | 27(47%) |
| BMI (Median) | 24.8 | 26.0 |
| Smoking n (%) | 12 (20.8%) | 13 (22.4%) |
| Co-morbidities n (%) |  |  |
| Diabetes | 4 (6.8%) | 0 (0%) |
| OSA | 1 (1.7%) | 0 (0%) |
| Hypertension | 2 (3.4%) | 0 (0%) |
| Cardiovascular Disease | 2 (3.4%) | 0 (0%) |
| Arthritis | 3 (5.1%) | 0 (0%) |
| Kidney | 2 (3.4%) | 0 (0%) |
| Asthma | 8 (13.7%) | 0 (0%) |
| Hyperlipidemia | 2 (3.4%) | 0 (0%) |
| Duration of infliximab combination therapy (median, months) | 12 | NA |
| Disease extent, n (%) |  |  |
| Ulcerative colitis (UC) | 24 (40%) | NA |
| E1: ulcerative proctitis | 4 (16.6) | NA |
| E2: left sided colitis | 7 (29.2%) | NA |
| E3: extensive colitis | 13 (54.2%) | NA |
| Crohn's disease (CD) | 34 (60%) | NA |
| L1: ileal | 17 (50%) | NA |
| L2: colonic | 4 (11.7%) | NA |
| L3: ileocolonic | 12 (35.2%) | NA |
| L4: upper gastrointestinal | 1 (3.1%) | NA |
| B1: inflammatory | 15 (44.1%) | NA |
| B2: stricturing | 9 (26.5%) | NA |
| B3: penetrating | 10 (29.4%) | NA |

\* *Infliximab combination therapy*

### Outcomes

Median (IQR) IgG were lower in patients treated with infliximab combination therapy [99 BAU/mL (40, 177)] than the healthy participants [139 BAU/mL (120, 188)], following vaccination with BNT162b2 (p = 0.0032). Neutralizing antibodies was also lower in the study group compared to control group [64 (23, 94) vs 91 (85, 94), p <0.001]. The median IgA levels in the study group was also significantly lower when compared to the control group [6 U/ml (3, 34) vs 13 U/ml (7, 30), p =0.0097]. (See table 2)

**Table 2:** Antibody response – Infliximab combination therapy (study group) vs healthy participants (control group)

| Characteristic | N | Overall, N = 116<sup>1</sup> | Study group, N = 58<sup>1</sup> | Control group, N = 58<sup>1</sup> | p-value<sup>2</sup> |
| --- | --- | --- | --- | --- | --- |
| **IgG BAU/mL** | 116 | 134 (86, 184) | 99 (40, 177) | 139 (120, 188) | 0.0032 |
| **IgA U/ml** | 116 | 9 (4, 30) | 6 (3, 34) | 13 (7, 30) | 0.0097 |
| **Neutralizing** | 116 | 87 (60, 94) | 64 (23, 94) | 91 (85, 94) | <0.001 |

<sup>1</sup>Median (IQR) or Frequency (%)
<sup>2</sup>Wilcoxon rank sum test

The percentage of patients who achieved positive IgG levels in the control group was 100%, whereas the percentage of patients of with positive IgG levels (>31.5 BAU/mL) in the study group was 81%. (Figure 1) The percentage of participants in the study group with positive neutralizing antibody level was 100%, whereas the percentage of patients in the study group was 75%. (Figure 2) Finally, the percentage of participants in the control group with positive IgA antibody level was 62%, whereas the percentage of patients in the study group was approximately 40%. (Figure3)

**Figure 1.**
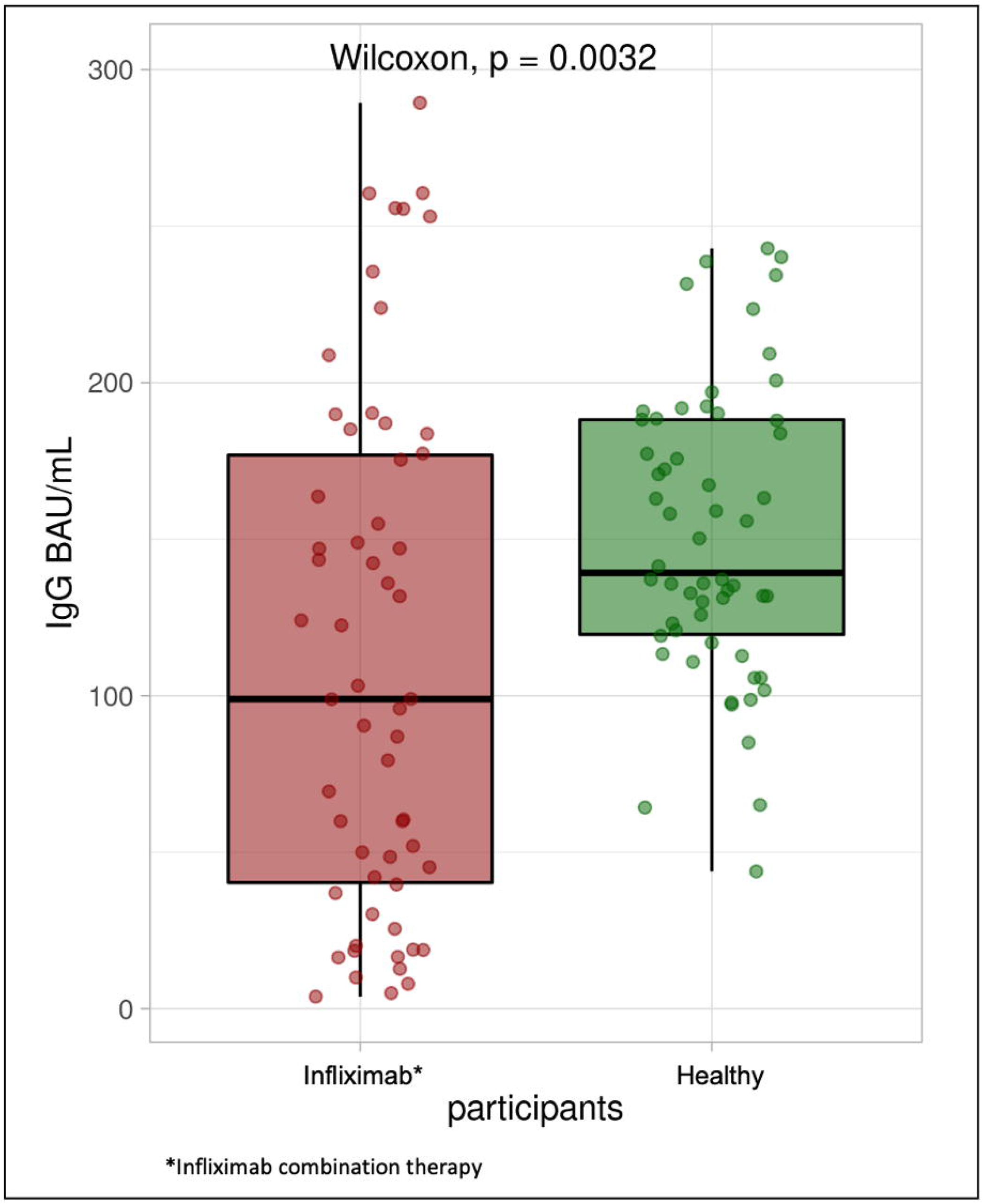
Box plot illustrating IgG antibody concentrations in the infliximab combination therapy group and healthy participants

**Figure 2.**
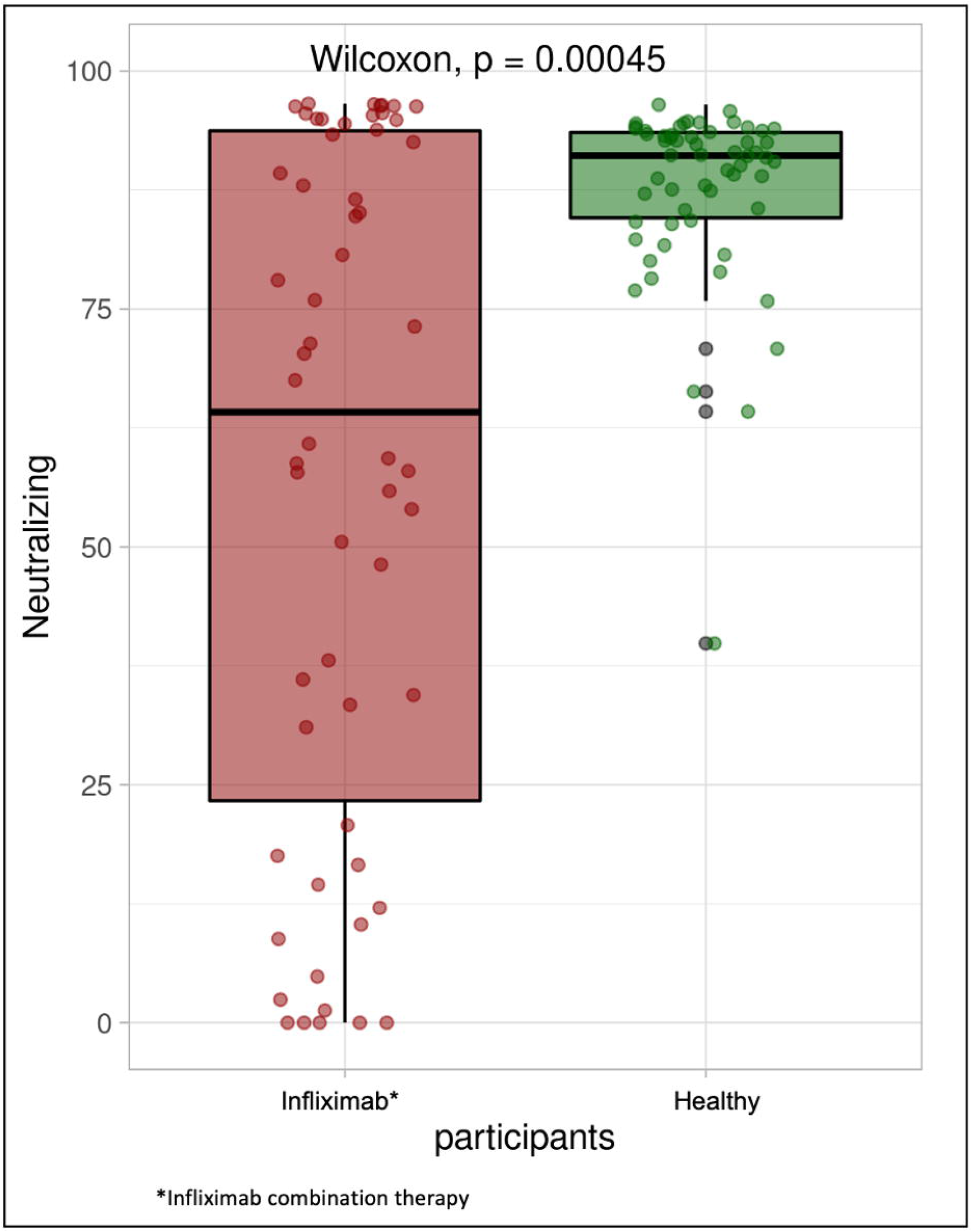
Box plot illustrating Neutralizing antibody concentrations in the infliximab combination therapy group and healthy participants

**Figure 3.**
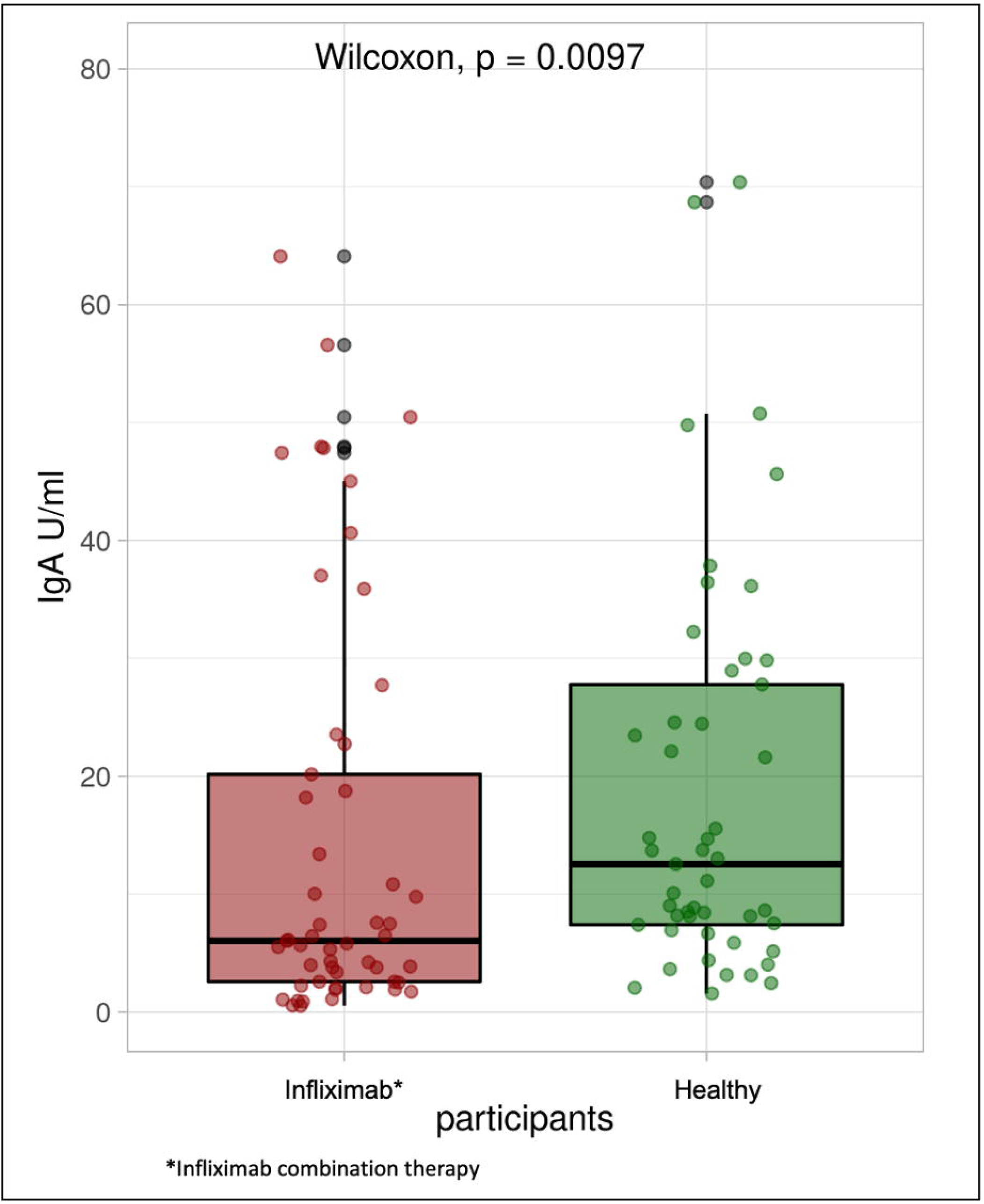
Box plot illustrating IgA antibody concentrations in the infliximab combination therapy group and healthy participants

## Discussion

In this study, we explored the serologic response to two doses of the BNT162b2 vaccine in IBD patients treated with infliximab combination therapy. We found that in patients with IBD receiving infliximab combination therapy, IgG, IgA and neutralizing antibody levels were lower compared to healthy participants 4-8 weeks following vaccination. However, the majority of patients treated with infliximab combination therapy achieved positive antibody concentrations consistent with those thought to provide protection against SARS-CoV-2 infection.

One study reported diminishing humoral responses within 6 months after receipt of the second dose of BNT162b2 vaccine in a large cohort of health care workers.^16^ However, another study observed a rapid decay in anti-SARS-CoV-2 antibodies, as early as 14 to 18 weeks, after completing the vaccination in patients with IBD receiving infliximab.^17^ Thus, in our study we only included patients who completed their second dose of vaccine within 4-8 weeks of recruitment.

One cohort study^18^ conducted in two centers in the United States and included 133 patients with inflammatory diseases and 53 immunocompetent patients found that 88.7% of patients with inflammatory disease achieved seroconversion after 2 doses of the BNT162b2 or mRNA-1273 vaccine. Similar to our study, the authors found that the antibody levels in patients with inflammatory disease were lower compared to the immunocompetent patients. They also observed that patients receiving corticosteroids had lower antibody titers after both vaccines. Similarly, in a cohort study of 300 patients, 24 patients were receiving infliximab combination therapy, and of those 88% achieved positive humoral immune response after complete vaccination with an mRNA vaccine.^19^

Two recent studies have explored the effect of immunosuppressive medications on serological response to SARS-CoV-2 infection and vaccination. Kennedy et al found that infliximab is correlated with diminished serological response to infection with SARS-CoV-2.^13^ The study also found that in patients receiving infliximab combination therapy, the serological response to infection was reduced further. Another study^20^ reported outcomes in patients with IBD receiving biological therapy after 2 dose vaccination of either Pfizer or Moderna vaccines. They found that for all antibodies tested, vedolizumab was associated with lower seropositivity, however, in patients receiving an anti-TNF agent lower antibody level was only for anti-receptor binding domain total Ig. Furthermore, a study conducted by Khan et al in veteran patients with IBD receiving different immunosuppressive medications found that among the 6578 patients with full vaccination status, no SARS-CoV-2 infections were identified among those taking anti-TNF agents, or ustekinumab.^21^ However, they reported that the level of protection in their IBD cohort was lower than that reported in the clinical trials with 80.4% effectiveness vs > 90%. Finally, a recent study^22^ investigated whether patients with inflammatory bowel disease treated with infliximab have attenuated serological responses to a single dose of Pfizer or Moderna vaccines. They concluded Infliximab is associated with lower seropositivity to a single dose of Pfizer or Moderna vaccines in patients with IBD. On the other hand, one study^23^ assessed antibody titers in adults with IBD who received mRNA SARS-CoV-2 vaccination who were referred from 18 U.S. gastroenterology practices. Participants were receiving various medication regimens including, anti-integrin therapy, anti–interleukin-12/23 therapy, and anti–tumor necrosis factor with or without immunomodulator. They found that 99% of participants had detectable antibodies after 2 weeks regardless of medication regimen.

Another important finding of this study is that while 75-80% of patients on infliximab combination therapy had positive IgG and neutralizing antibody levels, only 40% had positive IgA levels. SARS-CoV-2 circulating antibodies, particularly the IgG class, have been the major contributor to risk reduction of severe COVID-19 after vaccination.^24^ IgG antibodies specifically target the spike protein of SARS-CoV-2, its S1 subunit or its receptor binding domain (RBD). This diminish or completely halt the binding of the virus with the host receptors. Neutralizing antibodies have also been shown to correlate with protection.^25^ While most studies^26,27^ focused on the role if IgG or neutralizing antibodies in preventing severe COVID-19 illness after vaccination, the potential benefits of IgA antibodies in limiting infection and viral spread have been overlooked.^28^ Evidence is now accumulating regarding the IgA serum levels and its correlation to neutralization of SARS-CoV-2 at the mucosal surface.^29,30^ One study showed that IgA antibodies predominate the early neutralization of the virus and its appearance following infections seems earlier than that of antibodies of the IgG class.^31^ This study also observed a rapid decline in SARS-CoV-2–specific IgA serum levels, thereby bringing into question the long-term protective effect of IgA. This could be a possible explanation of the low level of IgA antibodies in our study as well as control group.

Our findings have relevant and immediate clinical implications. Our study supports the effectiveness of two dose vaccination in IBD patients receiving infliximab combination therapy. This can increase patient motivation to receive COVID-19 vaccine by demonstration and reassurance of vaccine effectiveness. In addition, we found that majority of patients on infliximab combination therapy achieved antibody levels that confers protection, while this prospect requires further study, it is not known whether higher antibody titer threshold correlates to protection against COVID-19 severe outcomes such as hospitalization and death. Therefore, British Society of Gastroenterology (BSG) and Food and Drug Administration (FDA) recommended a third dose (or booster dose) of SARS-CoV2 vaccine for all patients with IBD receiving immunosuppressive treatment.^32,33^ Furthermore, it remains to be seen whether infliximab combination therapy accelerates declining of titers over time, but our results may reassure patients receiving these medications that initial humoral responses to mRNA vaccines are generally robust. Future larger follow up studies needed.

To our knowledge this is the first prospective cohort study to compare the serological response of IBD patients on infliximab combination therapy to healthy participants. In addition, our predefined inclusion and exclusion criteria lowers the risk of confounding bias, and patients were equally distributed in terms of demographics characteristics such as age, gender and BMI.

One of the limitations of our study is a small sample size which may not reflect the precise difference between the two groups. Furthermore, even though we matched our participants in each group for known confounding factors, given the observational design, there is potential for unmeasured factors. In addition, we are only assed positive IgG, neutralizing antibody and IgA (humoral immunity). However, cellular immunity may also play a role in vaccine efficacy. Finally, we investigated infliximab with azathioprine or 6-mercaptopurine only. Further studies are needed to investigate the effect of other immunomodulators.

## Conclusion

In patients with IBD receiving infliximab combination therapy, IgG, IgA and neutralizing antibody levels were lower compared to healthy participants. However, the majority of patients treated with infliximab combination therapy achieved seropositivity after two doses of the vaccine.

## Supporting information

STROBE checklist

## Data Availability

All data produced in the present work are contained in the manuscript

## Funding statement

This Study was funded by Kuwait Foundation for the Advancement of Sciences (KFAS) grant (RA HM-2021-008).

## Notes

### Competing Interest Statement

The authors have declared no competing interest.

### Author Declarations

This study was reviewed and approved by the Ethical Review Board of Dasman institute (Protocol # RA HM-2021-008) as per the updated guidelines of the Declaration of Helsinki (64th WMA General Assembly, Fortaleza, Brazil, October 2013) and of the US Federal Policy for the Protection of Human Subjects. The study was also approved by the Ministry of Health of Kuwait (reference: 3799, protocol number 1729/2021). Subsequently, patient informed written consent was obtained before inclusion in the study.

